# Combined polygenic risk scores of different psychiatric traits predict general and specific psychopathology in childhood

**DOI:** 10.1101/2020.11.17.20233106

**Authors:** Alexander Neumann, Alexia Jolicoeur-Martineau, Eszter Szekely, Hannah M. Sallis, Kieran O’Donnel, Celia M.T. Greenwood, Robert Levitan, Michael J. Meaney, Ashley Wazana, Jonathan Evans, Henning Tiemeier

**Affiliations:** Department of Child and Adolescent Psychiatry, Erasmus University Medical Center Rotterdam, the Netherlands; Lady Davis Institute for Medical Research, Jewish General Hospital, Montreal, QC, Canada; VIB Center for Molecular Neurology, VIB, Antwerp, Belgium; Department of Biomedical Sciences, University of Antwerp, Antwerp, Belgium; Department of Psychiatry, McGill University Faculty of Medicine, Montreal, QC, Canada; MRC Integrative Epidemiology Unit, University of Bristol, Bristol, UK; Centre for Academic Mental Health, Population Health Sciences, Bristol Medical School, University of Bristol, Bristol, UK; School of Psychological Science, University of Bristol, Bristol, UK; Department of Psychiatry and Sackler Program for Epigenetics and Psychobiology, McGill University, Montreal, QC, Canada; Ludmer Centre for Neuroinformatics and Mental Health, McGill University, Montreal, QC, Canada; Departments of Oncology, Human Genetics, and Epidemiology, Biostatistics and Occupational Health, McGill University, Montreal, QC, Canada; Centre for Addiction and Mental Health, Toronto, ON, Canada; Department of Psychiatry, University of Toronto, Toronto, ON, Canada; Douglas Mental Health Institute, Montreal, QC, Canada; Singapore Institute for Clinical Sciences, Singapore City, Singapore; Centre for Child Development and Mental Health, Jewish General Hospital, Montreal, QC, Canada; Department of Social and Behavioral Sciences, Harvard T. H. Chan School of Public Health, Boston, MA, USA

**Keywords:** Genetics, molecular, Comorbidity, Internalising disorder, Externalising disorder, Meta-analysis

## Abstract

**Background:** Polygenic risk scores (PRSs) operationalize genetic propensity towards a particular mental disorder and hold promise as early predictors of psychopathology, but before a PRS can be used clinically, explanatory power must be increased and the specificity for a psychiatric domain established. To enable early detection it is crucial to study these psychometric properties in childhood. We examined whether PRSs associate more with general or with specific psychopathology in school-aged children. Additionally, we tested whether psychiatric PRSs can be combined into a multi-PRS score for improved performance.

**Methods:** We computed 16 PRSs based on GWASs of psychiatric phenotypes, but also neuroticism and cognitive ability, in mostly adult populations. Study participants were 9247 school-aged children from three population-based cohorts of the DREAM-BIG consortium: ALSPAC (UK), The Generation R Study (Netherlands) and MAVAN (Canada). We associated each PRS with general and specific psychopathology factors, derived from a bifactor model based on self-, parental-, teacher-, and observer reports. After fitting each PRS in separate models, we also tested a multi-PRS model, in which all PRSs are entered simultaneously as predictors of the general psychopathology factor.

**Results:** Seven PRSs were associated with the general psychopathology factor after multiple testing adjustment, two with specific externalizing and five with specific internalizing psychopathology. PRSs predicted general psychopathology independently of each other, with the exception of depression and depressive symptom PRSs. Most PRSs associated with a specific psychopathology domain, were also associated with general child psychopathology.

**Conclusions:** The results suggest that PRSs based on current GWASs of psychiatric phenotypes tend to be associated with general psychopathology, or both general and specific psychiatric domains, but not with one specific psychopathology domain only. Furthermore, PRSs can be combined to improve predictive ability. PRS users should therefore be conscious of non-specificity and consider using multiple PRSs simultaneously, when predicting psychiatric disorders.

## Introduction

Many psychiatric disorders have a strong genetic basis (Polderman et al., 2015), thus uncovering the genetic pathways underlying psychopathology holds the promise of individualized prediction and treatment. While most genome-wide associations studies (GWAS) of psychiatric disorders investigate distinct disorders, effects are often not unique to a specific disorder. For instance, GWAS-derived genetic correlations among psychiatric disorders average 0.41 (Anttila et al., 2018). Furthermore, a GWAS of eight disorders found 23 loci with strong evidence for association with at least four disorders (Lee et al., 2019).

The non-specificity of GWAS findings raises the issue of whether derived polygenic risk scores (PRS) can in fact predict specific psychiatric symptoms or disorders. PRSs are increasingly used in psychiatric research to operationalize the genetic predisposition towards a single disorder (Wray et al., 2020). However, before clinical adoption is entertained, it is crucial to understand what symptoms or disorders a given psychiatric PRS is in fact predicting.

Psychiatric symptoms are often grouped into internalizing (emotional problems like depression or anxiety) and externalizing problems (behavioral problems like aggression or conduct problems), but symptoms from both domains often co-occur, which has resulted in the development of hierarchical taxonomies.(Lahey, Moore, Kaczkurkin, & Zald, 2021) Within this taxonomy, psychiatric symptoms can be viewed as the result of a general psychopathology factor, underlying psychiatric symptoms from all domains, and more specific factors, which reflect the specific propensity towards one domain only. This hierarchical structure appears to be mirrored in the brain and genome (Sprooten, Franke, & Greven, 2021), but a comprehensive understanding of the specificity of PRSs for different domains is missing. This gap is even more pertinent in the context of child psychiatry, where symptoms are not as differentiated and often shift from one domain to another (Finsaas, Bufferd, Dougherty, Carlson, & Klein, 2018; Rutter, Kim-Cohen, & Maughan, 2006), e.g. children with ADHD developing depression in adolescence (Biederman et al., 2008). Genetic risks towards childhood psychiatric disorders are especially important to study the development of early prediction systems.

A few studies have evaluated the specificity of PRSs, e.g., a PRS of schizophrenia predicted post-traumatic stress, bipolar and anxiety disorders (Zheutlin et al., 2019). Furthermore, an ADHD PRS was more strongly associated with a general psychopathology factor, encompassing symptoms from multiple domains, than a specific ADHD factor (Brikell et al., 2018). In another study a principal component of eight different PRSs was associated with general psychopathology (Allegrini et al., 2019). However, an overview of different PRSs, which reports the degree to which they associate with general or specific psychopathology in childhood is missing.

In this study we examined i) whether individual PRSs derived from GWASs of specific psychiatric disorders, cognitive traits and neuroticism are predictive of general and/or specific externalizing and internalizing psychopathology in school-aged children; and, ii) the independent contribution of each PRS towards general psychopathology when combined with other PRSs. We hypothesized that—in addition to predicting their corresponding specific domain—each PRS is also associated with a general psychopathology factor. We further hypothesized that these associations will be substantially attenuated in mutually adjusted models. These hypotheses have been archived by the Generation R Data Management prior to analysis.

We studied these questions in the Developmental Research in Environmental Adversity, Mental health, BIological susceptibility and Gender (DREAM-BIG) project, a multi-center consortium of population-based cohorts with harmonized measures of psychopathology and genetics (Sallis et al., 2019; Szekely et al., 2020).

## Methods

### Participants

This study features three population-based prenatal cohorts: the Avon Longitudinal Study of Parents and Children (ALSPAC) from England (Boyd et al., 2013; Fraser et al., 2013), Generation R (GenR) from the Netherlands (Kooijman et al., 2016), and the Maternal Adversity, Vulnerability and Neurodevelopment (MAVAN) study from Canada (O’Donnell et al., 2014). Participants had information on at least one psychopathology subscale and on genotype. The ALSPAC Ethics and Law Committee, Medical Ethics Committee of the Erasmus Medical Center, Douglas Mental Health University Institute and St-Joseph’s Hospital approved the study. Only participants with European ancestry were included due to difficulties in applying PRSs derived from source GWAS of mostly European ancestry populations to other populations (Martin et al., 2019). One sibling per family was randomly excluded.

ALSPAC had 11,612 children with information on psychopathology, 6575 having genetic information. In GenR psychiatric information was available for 7946 children, 2418 were genotyped. MAVAN had 408 children with information on psychopathology, 254 had genetic information. All participants had European ancestry. The total sample size in meta-analyses was 9247 (Table 1, Supplemental Information 1).

**Table 1:** Cohort characteristics

| Characteristic | Category | ALSPAC | Category | GenR | Category | MAVAN |
| --- | --- | --- | --- | --- | --- | --- |
|  |  | M (SD)<br>or % |  | M (SD)<br>or % |  | M (SD)<br>or % |
| Sex | Female | 48.9% | Female | 51.0% | Female | 49.3% |
|  | Male | 51.1% | Male | 49.0% | Male | 50.7% |
| Avg. age at psychiatric assessments in years |  | 7.6<br>(0.2) |  | 6.6<br>(0.5) |  | 5.4<br>(0.1) |
| Maternal Age at Birth |  | 29.0<br>(4.6) |  | 31.7<br>(4.1) |  | 30.6<br>(4.9) |
| Maternal Education | CSE/None | 14.0% | None/Primary | 0.5% | Low | 14.4% |
|  | Vocational | 8.7% |  |  |  |  |
|  | O-level | 35.1% |  |  |  |  |
|  | A-level | 25.6% | Secondary | 28.1% | Medium | 33.5% |
|  | University | 16.6% | University | 71.5% | High | 52.1% |
| Income | Quintile 1 | 15.8% | <1,600€ p.m. | 5.1% | <15,000CAD p.y. | 5.6% |
|  | Quintile 2 | 18.7% | <2,400€ p.m. | 9.0% | <30,000CAD p.y. | 13.6% |
|  | Quintile 3 | 20.8% | <3,200€ p.m. | 18.5% | <45,000CAD p.y. | 11.3% |
|  | Quintile 4 | 21.8% | <4,800€ p.m. | 34.9% | <90,000CAD p.y. | 33.8% |
|  | Quintile 5 | 22.9% | ≥4,800€ p.m. | 32.5% | ≥ 90,000CAD p.y. | 35.7% |

**M** mean
**SD** Standard Deviation

### Measures

#### Polygenic Risk Scores Selection

We computed PRSs for 16 different psychiatric disorders, neuroticism and cognitive ability (Supplemental Information 1-2). We performed a systematic search of appropriate source GWAS on June 26^th^ 2019 by examining all GWAS listed in the psychiatric genetic consortium (PGC) data index (https://www.med.unc.edu/pgc/data-index/), in any consortia linked in the PGC data index (ANGST, Converge, Eagle, GPC, SSGAC, CCACE), and in the UK Biobank data fields “20544: Mental health problems ever diagnosed by a professional” and “1200: Sleeplessness/insomnia”. We further added an EAGLE GWAS on total psychiatric problems (Neumann et al., 2020).

#### Genotyping

Each cohort genotyped participants using SNP arrays and applied cohort-specific QC (Supplemental Information 1). Data was imputed to the HRC 1.1 reference panel using either the Michigan Imputation Server (Das et al., 2016) (ALSPAC and GenR) or Sanger Imputation Service (MAVAN) (McCarthy et al., 2016). SNPs with a minor allele frequency below 1% or imputation quality (R^2^) below 0.80 were excluded. In ALSPAC and GenR PRSs were calculated with PRSice 2 (Choi & O’Reilly, 2019) using default options (clumping correlated SNPs within a 250kb window at a r^2^ threshold of 0.1). In MAVAN an equivalent computation was performed with PRS-on-SPARK (PRSoS) using clumping setting of r^2^ = 0.25 within a window of 500kb (Chen et al., 2018). PRSice and PRSoS use equivalent method to compute PRSs, but differ in supported file formats and speed. All cohorts calculated PRSs at the following p-value thresholds:1,0.5,0.4,0.3,0.2,0.1,0.05,0.01,1×10-3,1×10-4,1×10-5,1×10-6,1×10-7,5×10-8,1×10-8. See Table S1-S3 for number of SNPs included.

#### Child Psychopathology

Each cohort had repeatedly collected several measures of psychopathology from 4 through 8 years of age. The average assessment age across instruments was 7.6 [range:7.5-8.3] years for ALSPAC, 6.6 [range:6.0-8.1] years for GenR and 5.4 [range:4.1-6.2] years for MAVAN. As children may behave differently in various environments (e.g. home vs school) and self-report at a young age is insufficient, we combined various instruments, including parental-, teacher-, self- and observer-rated, and diagnostic measures. Different assessment ages and informants were jointly modeled in each cohort to estimate factors representing consistently rated psychopathology levels in early school-age. See Sallis et al. (2019) and Supplemental Information for a complete description of instruments.

We estimated child psychopathology factors scores from a bifactor model (Sallis et al., 2019). Briefly, we used a bifactor model to define a general psychopathology factor, which underlies all measured psychopathology subscales, and two orthogonal specific internalizing and externalizing factors. These specific factors underlie the subscales of one domain only and represent internalizing or externalizing specific variance, which is not shared with the other domain or other psychopathology.

### Statistical Analysis

#### Separate PRS Models

We first analyzed the associations between each PRS and the three outcomes (general and specific externalizing/internalizing), separately. We regressed child psychopathology factors scores on each PRS at every threshold in separate regression models in each cohort. All analyses were adjusted for age, sex and ancestry (by including the first four components/dimensions of a genome-wide principal component analysis or multidimensional scaling).

Standardized regression coefficients and standard errors were extracted and meta-analyzed across cohorts. We applied a random-effect meta-analysis to account for potential sources of heterogeneity, e.g. different genotype and phenotype assessment methods and country differences. Specifically, we used the Han and Eskin method, which accounts for study heterogeneity, while retaining power comparable to fixed effects (Han & Eskin, 2011). We adjusted for multiple testing, by estimating the number of effective tests using the eigenvalues of the correlation matrix between all the PRSs and thresholds. We used the largest cohort (ALSPAC) to derive the correlation structure. According to the Li & Ji (2005) method, as implemented in poolR (Cinar & Viechtbauer), the number of independent tests is 99, resulting in a Bonferroni adjusted threshold of p<0.05/99<5.0×10^-4^. We express the variance explained as the difference in R^2^ compared to a covariates only model without PRSs. Additionally, we used sample-size weighted R values to compute an average R^2^ across cohorts. To test for differences in association strength between the specific factors and the general psychopathology factor, we applied z-tests (Clogg, Petkova, & Haritou, 1995).

#### Mutually Adjusted PRS Model

Next, we included all PRSs at their most significant threshold in a mutually adjusted PRS model to estimate the independent contribution of each PRS to general psychopathology in each cohort. More specifically, we fitted a regression model including all 16 PRSs, as predictors of the general psychopathology factor. From the PRSs of the same phenotype, only the threshold which showed the lowest p-value PRS was selected for the mutually adjusted analysis. The lowest p-value refers to the meta-analysis p-value across the three cohorts in the previous separate model, which could differ by outcome. Mutually adjusted PRS models were adjusted for the same covariates and meta-analyzed using the same approach as the separate PRS models. A PRS was considered to independently contribute to general psychopathology, if it showed multiple testing adjusted significance in separate PRS models and remained nominally significant in the mutually adjusted model (p<0.05). To quantify the variance in the general psychopathology jointly explained by PRSs, we applied a repeated (n=100) 10-fold repeated cross-validation in each cohort and subtracted by the variance explained by covariates only.

## Results

Seven PRSs were associated with general psychopathology in unadjusted models (Table 2, Figure 1), two PRSs were associated with the specific externalizing factor and five with the specific internalizing factor (Table 4, Figure 1). The PRSs for cognitive ability, ADHD, major depression, neuroticism, schizophrenia, insomnia and depressive symptoms were all associated with general psychopathology and explained between 0.17% and 0.99% of variance in general psychopathology (Table S1). Associations were in the expected directions, with a PRS for higher cognitive ability predicting lower general psychopathology, while a higher genetic risk for a psychiatric disorder or neuroticism was associated with a higher propensity for general psychopathology. Absolute effect sizes tended to be larger for PRSs with higher discovery sample sizes (Spearman’s ρ=0.38).

**Table 2:** General psychopathology factor regressed on PRS (each PRS in separate model)

| GWAS phenotype for PRS | PRS p cutoff | ALSPAC (n=6575) |  | GenR (n= 2418) |  | MAVAN (n=254) |  | Meta-Analysis (n=9247) |  |  |  |
| --- | --- | --- | --- | --- | --- | --- | --- | --- | --- | --- | --- |
| | | $\beta$ | SE | $\beta$ | SE | $\beta$ | SE | $\beta$ | $\tau$ | SE | p |
| Cognitive ability | 1 | -0.12 | 0.01 | -0.05 | 0.02 | -0.02 | 0.07 | -0.079 | 0.044 | 0.031 | 6.3E-25* |
| ADHD | 0.3 | 0.11 | 0.01 | 0.06 | 0.02 | 0.18 | 0.07 | 0.096 | 0.030 | 0.023 | 2.9E-23* |
| Major Depression | 0.05 | 0.07 | 0.01 | 0.04 | 0.02 | 0.11 | 0.07 | 0.060 | 0.000 | 0.010 | 2.2E-09* |
| Neuroticism | 0.01 | 0.05 | 0.01 | 0.07 | 0.02 | 0.13 | 0.07 | 0.056 | 0.000 | 0.010 | 3.0E-08* |
| Insomnia | 0.2 | 0.06 | 0.01 | 0.02 | 0.02 | 0.02 | 0.06 | 0.044 | 0.019 | 0.017 | 4.1E-07* |
| Schizophrenia | 0.05 | 0.05 | 0.01 | 0.05 | 0.02 | -0.07 | 0.18 | 0.051 | 0.000 | 0.010 | 6.7E-07* |
| Depressive symptoms | 0.3 | 0.05 | 0.01 | 0.00 | 0.02 | 0.07 | 0.07 | 0.032 | 0.032 | 0.024 | 4.3E-05* |
| Alcohol abuse | 0.2 | 0.04 | 0.01 | -0.03 | 0.02 | -0.03 | 0.07 | 0.005 | 0.042 | 0.030 | 2.4E-03 |
| Cross-disorder | 1 | 0.03 | 0.01 | 0.02 | 0.02 | 0.30 | 0.18 | 0.031 | 0.010 | 0.012 | 2.8E-03 |
| Bipolar | 0.0001 | 0.03 | 0.01 | 0.01 | 0.02 | 0.12 | 0.06 | 0.025 | 0.015 | 0.015 | 1.7E-02 |
| Autism | 0.1 | 0.00 | 0.01 | 0.07 | 0.02 | -0.04 | 0.07 | 0.021 | 0.049 | 0.033 | 1.1E-02 |
| Total problems | 0.2 | 0.02 | 0.01 | 0.02 | 0.02 | 0.05 | 0.10 | 0.022 | 0.000 | 0.010 | 3.4E-02 |
| Generalized anxiety | 0.01 | 0.02 | 0.01 | 0.03 | 0.02 | 0.01 | 0.07 | 0.020 | 0.000 | 0.010 | 5.4E-02 |
| Social anxiety | 0.01 | -0.02 | 0.01 | -0.01 | 0.02 | 0.08 | 0.06 | -0.013 | 0.015 | 0.015 | 9.1E-02 |
| Panic | 0.00001 | 0.00 | 0.01 | 0.04 | 0.02 | 0.07 | 0.06 | 0.021 | 0.018 | 0.016 | 1.7E-01 |
| Phobia | 0.2 | 0.01 | 0.01 | 0.02 | 0.02 | 0.04 | 0.07 | 0.014 | 0.000 | 0.010 | 1.8E-01 |

**PRS p cutoff** P-value of PRS threshold with most significant association with outcome
**$\beta$** Standardized regression coefficient in SD
**$\tau$** Random effect of study in SD
**SE** Standard Error
**p** P-value of regression coefficient
\* Significant p-value after multiple testing adjustment ( $p < 5.0E-04$ )
**Note:** Standard errors for the schizophrenia, cross-disorder (which includes schizophrenia) and bipolar PRS were inflated in the MAVAN cohort due to multicollinearity with genetic ancestry. Estimates are unbiased, but have higher uncertainty. Uncertainty is taken into account in the pooled estimates.

**Figure 1:**
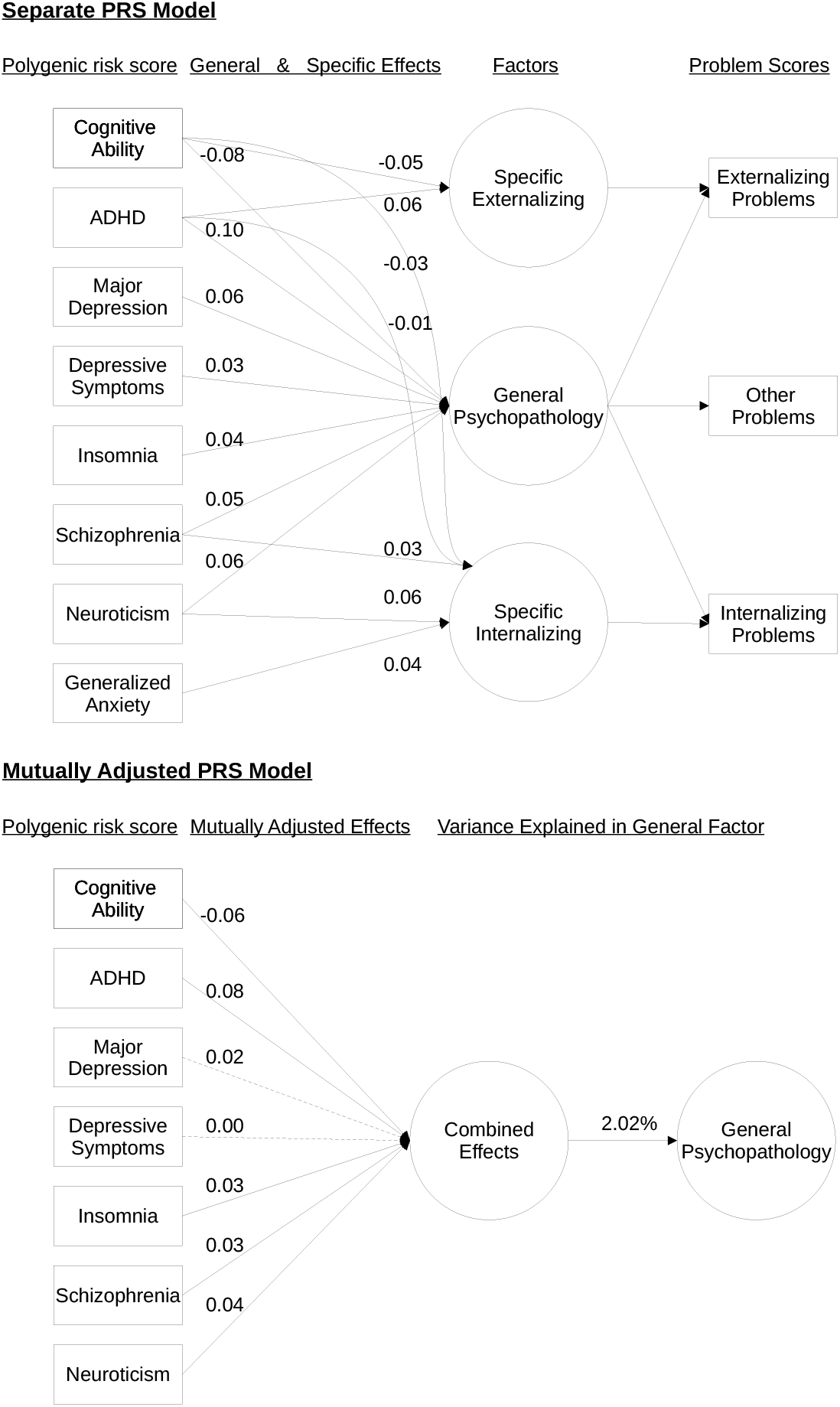
PRS model summary. General and specific psychopathology factors were regressed on 16 different PRSs. Only paths showing a significant association in separate PRS models after multiple testing adjustment (p<5.0E-04) are displayed and their corresponding standardized regression estimates. Dashed lines indicate a p-value of >0.05 in mutually adjusted model.

All PRSs correlated only modestly at their optimal threshold (r<0.34) (Figure S2). When modeling all PRS phenotypes jointly in a mutually adjusted model, the seven PRSs showing an association in separate PRS models also showed contributions to general psychopathology independent of each other (Table 3, Figure 1), with the exception of major depression and depressive symptoms. These seven PRSs jointly explained 2.02% of general psychopathology variance based on the average cross-validated performance across cohorts. Including all PRSs did not further improve performance (ΔR^2^ = 1.94%). The lack of independent association for the PRSs for major depression and depressive symptoms was not explained by the inclusion of two depression-related PRS in the model but rather by the inclusion of non-depression PRSs (Supplemental Information 1).

**Table 3:** General psychopathology factor regressed on PRS (mutually adjusted PRS model)

| GWAS phenotype for PRS | PRS p cutoff | ALSPAC (n=6575) |  | GenR (n= 2418) |  | MAVAN (n=254) |  | Meta-Analysis (n=9247) |  |  |  |
| --- | --- | --- | --- | --- | --- | --- | --- | --- | --- | --- | --- |
| | | $\beta$ | SE | $\beta$ | SE | $\beta$ | SE | $\beta$ | $\tau$ | SE | p |
| Cognitive ability | 1 | -0.09 | 0.01 | -0.04 | 0.02 | 0.03 | 0.08 | -0.059 | 0.038 | 0.027 | 3.8E-15* |
| ADHD | 0.3 | 0.10 | 0.01 | 0.04 | 0.02 | 0.19 | 0.08 | 0.084 | 0.039 | 0.028 | 6.4E-17* |
| Major Depression | 0.05 | 0.02 | 0.01 | 0.00 | 0.02 | 0.05 | 0.08 | 0.019 | 0.000 | 0.011 | 9.6E-02 |
| Neuroticism | 0.01 | 0.02 | 0.01 | 0.06 | 0.02 | 0.12 | 0.08 | 0.040 | 0.025 | 0.021 | 7.6E-03* |
| Insomnia | 0.2 | 0.04 | 0.01 | 0.00 | 0.02 | -0.01 | 0.07 | 0.026 | 0.010 | 0.012 | 7.5E-03* |
| Schizophrenia | 0.05 | 0.03 | 0.01 | 0.03 | 0.02 | -0.16 | 0.19 | 0.033 | 0.000 | 0.010 | 2.1E-03* |
| Depressive symptoms | 0.3 | 0.02 | 0.01 | -0.03 | 0.02 | 0.03 | 0.08 | 0.003 | 0.029 | 0.022 | 2.0E-01 |
| Alcohol abuse | 0.2 | 0.02 | 0.01 | -0.03 | 0.02 | -0.05 | 0.07 | -0.008 | 0.034 | 0.025 | 1.5E-01 |
| Cross-disorder | 1 | 0.00 | 0.01 | -0.01 | 0.02 | 0.20 | 0.19 | -0.002 | 0.000 | 0.010 | 8.9E-01 |
| Bipolar | 0.0001 | 0.01 | 0.01 | 0.00 | 0.02 | 0.09 | 0.07 | 0.010 | 0.000 | 0.010 | 3.7E-01 |
| Autism | 0.1 | -0.03 | 0.01 | 0.05 | 0.02 | -0.10 | 0.07 | -0.010 | 0.053 | 0.036 | 1.2E-02 |
| Total problems | 0.2 | 0.01 | 0.01 | 0.01 | 0.02 | -0.02 | 0.10 | 0.009 | 0.000 | 0.010 | 4.0E-01 |
| Generalized anxiety | 0.01 | 0.00 | 0.01 | 0.02 | 0.02 | -0.01 | 0.08 | 0.007 | 0.000 | 0.010 | 5.2E-01 |
| Social anxiety | 0.01 | -0.03 | 0.01 | -0.01 | 0.02 | 0.08 | 0.07 | -0.019 | 0.013 | 0.014 | 2.6E-02 |
| Panic | 0.00001 | 0.01 | 0.01 | 0.04 | 0.02 | 0.10 | 0.06 | 0.026 | 0.019 | 0.017 | 6.4E-02 |
| Phobia | 0.2 | 0.01 | 0.01 | 0.02 | 0.02 | -0.02 | 0.07 | 0.010 | 0.000 | 0.010 | 3.7E-01 |

**PRS p cutoff** P-value of PRS threshold with most significant association with outcome
**$\beta$** Standardized regression coefficient in SD
**$\tau$** Random effect of study in SD
**SE** Standard Error
**p** P-value of regression coefficient
\* PRS with nominal significance in mutually adjusted model and multiple testing adjusted significance in separate PRS models

For the specific externalizing psychopathology, only the ADHD and cognitive ability PRSs contributed robustly (Table 4). A genetic predisposition towards ADHD and lower cognitive ability was less predictive of specific externalizing psychopathology than of general psychopathology, explaining 0.13-0.15% variance (Table S2).

**Table 4:**
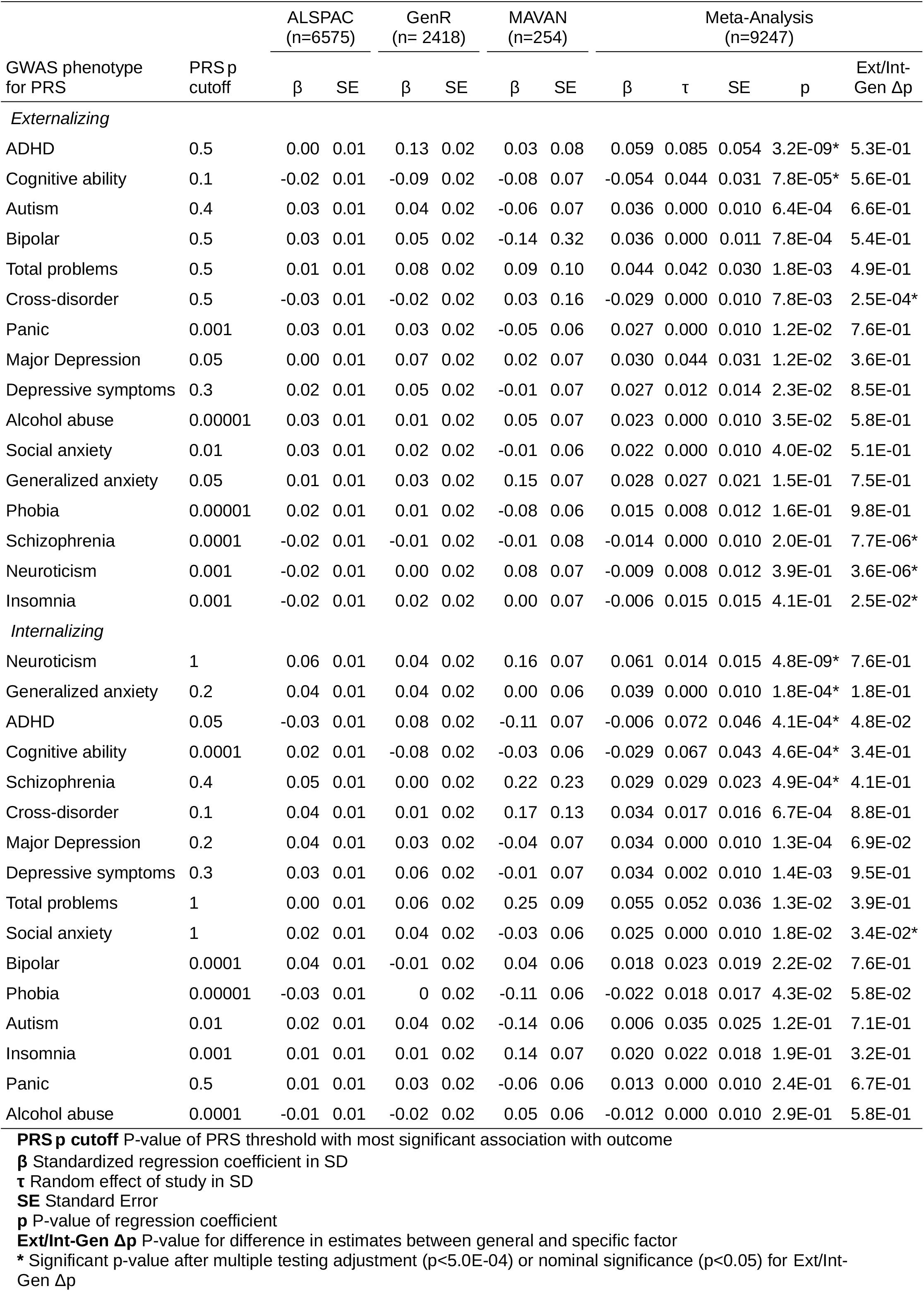
Specific psychopathology factors regressed on PRS (separate PRS model)

| GWAS phenotype for PRS | PRS p cutoff | ALSPAC (n=6575) |  | GenR (n= 2418) |  | MAVAN (n=254) |  | Meta-Analysis (n=9247) |  |  |  | Ext/Int-Gen Δp |
| --- | --- | --- | --- | --- | --- | --- | --- | --- | --- | --- | --- | --- |
|  |  | β | SE | β | SE | β | SE | β | τ | SE | p |  |
| Externalizing |  |  |  |  |  |  |  |  |  |  |  |  |
| ADHD | 0.5 | 0.00 | 0.01 | 0.13 | 0.02 | 0.03 | 0.08 | 0.059 | 0.085 | 0.054 | 3.2E-09* | 5.3E-01 |
| Cognitive ability | 0.1 | -0.02 | 0.01 | -0.09 | 0.02 | -0.08 | 0.07 | -0.054 | 0.044 | 0.031 | 7.8E-05* | 5.6E-01 |
| Autism | 0.4 | 0.03 | 0.01 | 0.04 | 0.02 | -0.06 | 0.07 | 0.036 | 0.000 | 0.010 | 6.4E-04 | 6.6E-01 |
| Bipolar | 0.5 | 0.03 | 0.01 | 0.05 | 0.02 | -0.14 | 0.32 | 0.036 | 0.000 | 0.011 | 7.8E-04 | 5.4E-01 |
| Total problems | 0.5 | 0.01 | 0.01 | 0.08 | 0.02 | 0.09 | 0.10 | 0.044 | 0.042 | 0.030 | 1.8E-03 | 4.9E-01 |
| Cross-disorder | 0.5 | -0.03 | 0.01 | -0.02 | 0.02 | 0.03 | 0.16 | -0.029 | 0.000 | 0.010 | 7.8E-03 | 2.5E-04* |
| Panic | 0.001 | 0.03 | 0.01 | 0.03 | 0.02 | -0.05 | 0.06 | 0.027 | 0.000 | 0.010 | 1.2E-02 | 7.6E-01 |
| Major Depression | 0.05 | 0.00 | 0.01 | 0.07 | 0.02 | 0.02 | 0.07 | 0.030 | 0.044 | 0.031 | 1.2E-02 | 3.6E-01 |
| Depressive symptoms | 0.3 | 0.02 | 0.01 | 0.05 | 0.02 | -0.01 | 0.07 | 0.027 | 0.012 | 0.014 | 2.3E-02 | 8.5E-01 |
| Alcohol abuse | 0.00001 | 0.03 | 0.01 | 0.01 | 0.02 | 0.05 | 0.07 | 0.023 | 0.000 | 0.010 | 3.5E-02 | 5.8E-01 |
| Social anxiety | 0.01 | 0.03 | 0.01 | 0.02 | 0.02 | -0.01 | 0.06 | 0.022 | 0.000 | 0.010 | 4.0E-02 | 5.1E-01 |
| Generalized anxiety | 0.05 | 0.01 | 0.01 | 0.03 | 0.02 | 0.15 | 0.07 | 0.028 | 0.027 | 0.021 | 1.5E-01 | 7.5E-01 |
| Phobia | 0.00001 | 0.02 | 0.01 | 0.01 | 0.02 | -0.08 | 0.06 | 0.015 | 0.008 | 0.012 | 1.6E-01 | 9.8E-01 |
| Schizophrenia | 0.0001 | -0.02 | 0.01 | -0.01 | 0.02 | -0.01 | 0.08 | -0.014 | 0.000 | 0.010 | 2.0E-01 | 7.7E-06* |
| Neuroticism | 0.001 | -0.02 | 0.01 | 0.00 | 0.02 | 0.08 | 0.07 | -0.009 | 0.008 | 0.012 | 3.9E-01 | 3.6E-06* |
| Insomnia | 0.001 | -0.02 | 0.01 | 0.02 | 0.02 | 0.00 | 0.07 | -0.006 | 0.015 | 0.015 | 4.1E-01 | 2.5E-02* |
| Internalizing |  |  |  |  |  |  |  |  |  |  |  |  |
| Neuroticism | 1 | 0.06 | 0.01 | 0.04 | 0.02 | 0.16 | 0.07 | 0.061 | 0.014 | 0.015 | 4.8E-09* | 7.6E-01 |
| Generalized anxiety | 0.2 | 0.04 | 0.01 | 0.04 | 0.02 | 0.00 | 0.06 | 0.039 | 0.000 | 0.010 | 1.8E-04* | 1.8E-01 |
| ADHD | 0.05 | -0.03 | 0.01 | 0.08 | 0.02 | -0.11 | 0.07 | -0.006 | 0.072 | 0.046 | 4.1E-04* | 4.8E-02 |
| Cognitive ability | 0.0001 | 0.02 | 0.01 | -0.08 | 0.02 | -0.03 | 0.06 | -0.029 | 0.067 | 0.043 | 4.6E-04* | 3.4E-01 |
| Schizophrenia | 0.4 | 0.05 | 0.01 | 0.00 | 0.02 | 0.22 | 0.23 | 0.029 | 0.029 | 0.023 | 4.9E-04* | 4.1E-01 |
| Cross-disorder | 0.1 | 0.04 | 0.01 | 0.01 | 0.02 | 0.17 | 0.13 | 0.034 | 0.017 | 0.016 | 6.7E-04 | 8.8E-01 |
| Major Depression | 0.2 | 0.04 | 0.01 | 0.03 | 0.02 | -0.04 | 0.07 | 0.034 | 0.000 | 0.010 | 1.3E-04 | 6.9E-02 |
| Depressive symptoms | 0.3 | 0.03 | 0.01 | 0.06 | 0.02 | -0.01 | 0.07 | 0.034 | 0.002 | 0.010 | 1.4E-03 | 9.5E-01 |
| Total problems | 1 | 0.00 | 0.01 | 0.06 | 0.02 | 0.25 | 0.09 | 0.055 | 0.052 | 0.036 | 1.3E-02 | 3.9E-01 |
| Social anxiety | 1 | 0.02 | 0.01 | 0.04 | 0.02 | -0.03 | 0.06 | 0.025 | 0.000 | 0.010 | 1.8E-02 | 3.4E-02* |
| Bipolar | 0.0001 | 0.04 | 0.01 | -0.01 | 0.02 | 0.04 | 0.06 | 0.018 | 0.023 | 0.019 | 2.2E-02 | 7.6E-01 |
| Phobia | 0.00001 | -0.03 | 0.01 | 0 | 0.02 | -0.11 | 0.06 | -0.022 | 0.018 | 0.017 | 4.3E-02 | 5.8E-02 |
| Autism | 0.01 | 0.02 | 0.01 | 0.04 | 0.02 | -0.14 | 0.06 | 0.006 | 0.035 | 0.025 | 1.2E-01 | 7.1E-01 |
| Insomnia | 0.001 | 0.01 | 0.01 | 0.01 | 0.02 | 0.14 | 0.07 | 0.020 | 0.022 | 0.018 | 1.9E-01 | 3.2E-01 |
| Panic | 0.5 | 0.01 | 0.01 | 0.03 | 0.02 | -0.06 | 0.06 | 0.013 | 0.000 | 0.010 | 2.4E-01 | 6.7E-01 |
| Alcohol abuse | 0.0001 | -0.01 | 0.01 | -0.02 | 0.02 | 0.05 | 0.06 | -0.012 | 0.000 | 0.010 | 2.9E-01 | 5.8E-01 |

**PRS p cutoff** P-value of PRS threshold with most significant association with outcome
**$\beta$** Standardized regression coefficient in SD
**$\tau$** Random effect of study in SD
**SE** Standard Error
**p** P-value of regression coefficient
**Ext/Int-Gen $\Delta p$** P-value for difference in estimates between general and specific factor
\* Significant p-value after multiple testing adjustment ( $p < 5.0E-04$ ) or nominal significance ( $p < 0.05$ ) for Ext/Int-Gen $\Delta p$

For specific internalizing psychopathology, we observed associations with the PRSs for neuroticism, generalized anxiety, ADHD, cognitive ability and schizophrenia (Table 4, S3). The effect size of the neuroticism PRS was similar for the specific internalizing factor and for general psychopathology, larger for the generalized anxiety PRS, and lower for ADHD, cognitive ability and schizophrenia. The explained variance ranged from 0.14-0.38% (Table S3). The pooled effect for the ADHD PRS was near 0, but nevertheless significant due to a very robust association in GenR, which influences the random-effects p-value. It should be noted, that the evidence for effect size difference between general and specific psychopathology was weak for all effect size comparisons.

## Discussion

Several PRSs associated with general and specific internalizing/externalizing psychopathology in children across three independent cohorts. Seven PRSs, representing the genetic propensity towards cognitive ability, ADHD, major depression, neuroticism, schizophrenia, insomnia and depressive symptoms, were associated with general psychopathology in school-aged children. All but two (major depression and depressive symptoms) PRSs contributed independently towards general psychopathology. Two PRSs were associated with specific externalizing psychopathology: ADHD and cognitive ability. Five PRSs were associated with specific internalizing psychopathology: neuroticism, generalized anxiety, ADHD, cognitive ability and schizophrenia. In general, the PRS associations support the validity of the bifactor structure of child psychopathology, with genetic predictors from various psychiatric domains being associated with general psychopathology and a narrower, more domain specific set of PRSs associating with specific psychopathology.

The main finding of this study is that PRSs for psychiatric and psychological traits are unlikely to be associated with domain-specific psychopathology exclusively in childhood. PRSs associated with school-age psychopathology tended to either associate with general psychopathology only, or both general and specific psychopathology, but not with specific psychopathology only, with the exception of generalized anxiety. Brikell et al. (2018) demonstrated this previously for an ADHD PRS and we confirm this to be a more general trend for psychiatric PRSs. It follows, that a PRS based on the GWAS of a specific psychiatric disorder may be a good predictor for that disorder, but is also likely to be predictive of other psychiatric domains. In fact, effect sizes tended to be larger for general than specific psychopathology. This may indicate that PRSs for psychiatric disorders heavily weigh SNPs with cross-disorder effects in childhood. On the one hand, this reflects the comorbid nature of psychiatric disorders. On the other hand, this makes interpretation of PRS associations difficult. A child scoring high on a PRS of a specific psychiatric disorder could actually develop many distinct symptoms from different domains. The development of more specific PRSs in combination of general PRSs are therefore needed for more complete projections of symptom profiles. Such PRSs could be potentially obtained from general psychopathology adjusted GWASs. Until then, researchers and clinicians must take these cross-phenotype associations into account when interpreting PRS results. Caution is especially warranted when using a PRS as genetic instrument for specific disorders or symptoms in Mendelian randomization studies. Most PRSs would likely violate the exclusion assumption, i.e. they may affect the outcome via pathways that do not involve the specific disorder they were computed to predict.

Curiously, a PRS for ADHD was associated with both specific externalizing and internalizing factors. It is possible that the ADHD GWAS also captures disruptive mood dysregulation disorder, which is characterized by the occurrence of key internalizing and externalizing traits. However, we need to cautiously interpret this finding, as it was very inconsistent between cohorts.

Another implication of the results is that in the pursuit of improving genetic predictions of psychiatric disorders, researchers should not only consider computing PRSs based on GWAS of the trait they intend to predict, but also consider related traits. As example, cognitive ability was one of the best predictors of general and specific psychopathology. This does not mean that cognition-related SNPs are more strongly associated with general psychopathology than psychiatric SNPs. Rather, the robust association may partly be explained by the large sample size of the discovery GWAS. Thus, PRSs of related traits may be especially useful, when large source GWAS of the target trait are lacking.

Most PRSs associated with general psychopathology had unique effects. Thus, the third implication of our study is that multiple PRSs should be used jointly for improved prediction of general psychopathology. However, currently the inclusion of depression PRSs may be redundant, as SNPs included in the depression PRSs with general effects can be expressed quite well as linear combination of general effects from other PRSs. The joint model explained twice the variance in general psychopathology factor than the most predictive PRSs alone. The findings support the notion, that combining information from different discovery GWASs improves prediction, which has been demonstrated before for cognition (Krapohl et al., 2018) and adult psychiatric disorders (Maier et al., 2015). Allegrini et al., (2019) explained almost 1% variance in general psychopathology in childhood based on a combination of eight PRSs. The estimated variance explained was approximately 2% in this study, possibly due to the inclusion of a PRS for cognitive ability.

A strength of this study was the prospective meta-analysis approach. Our study is the first attempt to harmonize genetic risk scores and latent constructs of child psychopathology in multiple independent cohorts. We benefited in particular from the inclusion of a wide range of measures, the inclusion of repeated assessments and multiple informants. Besides the improved precision through increased sample size, we also expect the results to generalize better to other populations of European ancestry compared to a single cohort study. Further investigations are needed in non-European ancestry populations to determine to what extent the results generalize, or whether predictions are attenuated. Another strength is the systematic search and selection of PRSs. This enabled us to test a wide variety of PRSs and form conclusions based on the current state of psychiatric PRSs as a whole.

Limitations of the study were pre-processing and PRS analysis pipelines differences between cohorts, which lead to different SNP sets being used in the calculation of PRSs. Furthermore, while the average assessment ages were fairly consistent between cohorts, they did range from four to eight years. Genetic effects tend to increase with age for most psychiatric disorders and thus age-specific effects should be explored in future studies. (Bergen, Gardner, & Kendler, 2007) Effect size differences between general and specific pathways tended to be smaller than the absolute effect size itself, thus the power to detect these differences was likely limited. The explained variance in general psychopathology (ca. 2%) is improved by inclusion of multiple PRSs, but remains much lower than the estimated SNP heritability of 18 to 36% (Alnæs et al., 2018; Neumann et al., 2016). The low explained variance likely limits the clinical utility of the multi-PRS score to detection of extreme genetic predisposition towards general psychopathology in childhood. The use of more childhood-specific discovery GWASs may help close this performance gap, but these tend to have much lower sample sizes than adulthood-based GWASs. As PRS performance is directly tied to the sample size of the discovery GWAS, the field relies on GWAS of adult-based GWASs (Raffington, Mallard, & Harden, 2020). However, the insights gained from this study in regard to specificity and independence of PRSs will hopefully in combination with better powered source GWAS help in the development of multi-PRS scores with high explanatory power and clinical utility.

In conclusion, our findings demonstrate that many PRSs for psychiatric traits are associated with general psychopathology in school-aged children. These effects were mostly independent of each other with the exception of depression-related PRS effects. Several PRSs were associated with general psychopathology and also specific externalizing or internalizing psychopathology, but only one PRS (generalized anxiety) was exclusively associated with specific internalizing psychopathology without being associated with general psychopathology. Finally, we recommend that researchers should use a combination of multiple PRSs if they want to improve prediction of child psychiatric symptoms.

## Data Availability

To ensure participant privacy and law compliance, individual-level data cannot be made publicly available without explicit informed consent, which is not available. For new analyses or individual-level data access, please contact Generation R data management and the corresponding author.

https://generationr.nl/researchers/collaboration/

http://www.bristol.ac.uk/alspac/researchers/access/

http://dreambigresearch.com/

## ACKNOWLEDGMENTS

ALSPAC

We are extremely grateful to all the families who took part in this study, the midwives for their help in recruiting them, and the whole ALSPAC team, which includes interviewers, computer and laboratory technicians, clerical workers, research scientists, volunteers, managers, receptionists and nurses. The UK Medical Research Council MRC and Wellcome (Grant ref: 217065/Z/19/Z) and the University of Bristol provide core support for ALSPAC, see http://www.bristol.ac.uk/alspac/external/documents/grant-acknowledgements.pdf. This publication is the work of the authors, who serve as guarantors for the contents of this paper. GWAS data was generated by Sample Logistics and Genotyping Facilities at Wellcome Sanger Institute and LabCorp (Laboratory Corporation of America) using support from 23andMe. This work was supported by the MRC and the University of Bristol (MC_UU_00011/7). H.M.Sallis is also supported by the European Research Council (758813 MHINT).

GenR

The Generation R Study is conducted by the Erasmus Medical Center in close collaboration with the Erasmus University Rotterdam, Faculty of Social Sciences, the Municipal Health Service Rotterdam area, and the Stichting Trombosedienst and Artsenlaboratorium Rijnmond. We gratefully acknowledge the contribution of general practitioners, hospitals, midwives and pharmacies in Rotterdam. GenR is financially supported by Erasmus Medical Center and the Netherlands Organization for Health Research and Development. A. Neumann and H. Tiemeier are supported by a grant of the Dutch Ministry of Education, Culture, and Science and the Netherlands Organization for Scientific Research (024.001.003, Consortium on Individual Development). The work of H. Tiemeier is further supported by a European Union’s Horizon 2020 research and innovation program (633595, DynaHealth) and a NWO-VICI grant (016.VICI.170.200).

MAVAN

The MAVAN Research Team sincerely thanks the families that have participated in the MAVAN project for so generously giving their time as well as their ongoing support. MAVAN was funded by a Canadian Institutes of Health Research (CIHR) trajectory grant (191827). Private support was received from the McGill University Faculty of Medicine, the Norlien Foundation, and the Woco Foundation. This study was made possible by the CHIR (359912, 365309, and 231614), the Fonds de la recherche en santé du Québec (22418), and the March of Dimes Foundation (12-FY12-198). R. Levitan is supported by the Cameron Parker Holcombe Wilson Chair in Depression Studies, University of Toronto and the Centre for Addiction and Mental Health.

The authors have declared that they have no competing/potential conflicts of interest.

## ABBREVIATIONS

PRS: Polygenic Risk Score
GWAS: Genome-Wide Association Study
SNP: Single Nucleotide Polymorphism
DREAM-BIG: Developmental Research in Environmental Adversity, Mental health, Biological susceptibility and Gender
ALSPAC: Avon Longitudinal Study of Parents and Children
GenR: Generation R
MAVAN: Maternal Adversity, Vulnerability and Neurodevelopment

## Keywords

- Polygenic risk scores (PRS) quantify a genetic predisposition towards a psychiatric disorder
- The specificity of most PRSs in predicting child psychiatric problems is unknown, i.e. it is unclear whether PRSs are particularly associated with specific symptoms or general psychopathology
- We systematically searched for GWAS of psychiatric or related phenotypes, and computed 16 PRSs in three cohorts (n = 9,247 school-aged children)
- PRSs tended to associate with general psychopathology only, or with both general and specific psychopathology, but not with specific psychopathology only
- Associations of different PRSs were mostly independent of each other, suggesting that different PRSs must be combined to predict childhood psychopathology in research and clinical practice

