## Supplemental Information 1 for "Combined polygenic risk scores of different psychiatric traits predict general and specific psychopathology in childhood"

Additional notes for methodology

#### Participants

##### ALSPAC

Pregnant women resident in Avon, UK with expected dates of delivery 1st April 1991 to 31st December 1992 were invited to take part in the the Avon Longitudinal Study of Parents and Children (ALSPAC). The initial number of pregnancies enrolled is 14,541 (for these at least one questionnaire has been returned or a “Children in Focus” clinic had been attended by 19/07/99). Of these initial pregnancies, there was a total of 14,676 foetuses, resulting in 14,062 live births and 13,988 children who were alive at 1 year of age. When the oldest children were approximately 7 years of age, an attempt was made to bolster the initial sample with eligible cases who had failed to join the study originally. As a result, when considering variables collected from the age of seven onwards (and potentially abstracted from obstetric notes) there are data available for more than the 14,541 pregnancies mentioned above. The number of new pregnancies not in the initial sample (known as Phase I enrolment) that are currently represented on the built files and reflecting enrolment status at the age of 24 is 913 (456, 262 and 195 recruited during Phases II, III and IV respectively), resulting in an additional 913 children being enrolled. The phases of enrolment are described in more detail in the cohort profile paper and its update. The total sample size for analyses using any data collected after the age of seven is therefore 15,454 pregnancies, resulting in 15,589 foetuses. Of these 14,901 were alive at 1 year of age.

Please note that the study website contains details of all the data that is available through a fully searchable data dictionary and variable search tool (http://www.bristol.ac.uk/alspac/researchers/our-data/).

Ethical approval for the study was obtained from the ALSPAC Ethics and Law Committee and the Local Research Ethics Committees. Informed consent for the use of data collected via questionnaires and clinics was obtained from participants following the recommendations of the ALSPAC Ethics and Law Committee at the time. Consent for biological samples has been collected in accordance with the Human Tissue Act (2004).

### PRS selection

As ALSPAC and GenR contributed to the EAGLE GWAS, the meta-analysis for the EAGLE GWAS was reran without these two cohorts to avoid overfitting. A GWAS result was retained for consideration if the study phenotype was a psychiatric diagnosis or continuous psychiatric symptom score, a combination of disorders, a measure of neuroticism or cognitive ability. Exclusion criteria included: GWASs of single symptoms; GWASs with insufficient precision of estimates, ie with fewer than 30,000 subjects, or case-control studies with fewer than 1000 cases. For each phenotype, the GWAS with the largest sample size or number of cases was retained. Major depression and continuously scaled depressive symptoms were treated as separate phenotypess, given the modest correlation between these two PRSs (0.38 between PRS including all SNPs). As the GWAS results for combined schizophrenia and bipolar disorder correlated strongly (r>0.90) with the original separate schizophrenia and bipolar GWASs, we used the separate PRS for bipolar and schizophrenia to avoid redundancy. The final selection consisted of 16 polygenic risk scores.

Despite similar QC criteria, SNPs can pass QC in some cohorts but not others, which can lead to the inability to compute some PRS at very strict p-value thresholds with few SNPs. We excluded PRS thresholds, which were not available in all three cohorts.

#### Genotyping

ALSPAC children were genotyped using the Illumina HumanHap550 quad chip genotyping platforms. Individuals were excluded on the basis of gender mismatches; minimal or excessive heterozygosity; disproportionate levels of individual missingness (>3%), insufficient sample replication (IBD < 0.8) or non-European ancestry. SNPs with a minor allele frequency of < 1%, a call rate of < 95% or evidence for violations of Hardy-Weinberg equilibrium (P < 5E-7) were removed. After combining genotype data in the mothers and the children, SNPs with genotype missingness above 1% or potential ID mismatches were removed.

For GenR, Illumina 610K/660W chips were used to analyze DNA from whole blood. QC filters included sample (≥97.5%) and SNP call rates (≥95%), minor allele frequency ≥1% and Hardy-Weinberg equilibrium deviations ($p<10^{-7}$). We tested excess heterozygosity, gender accuracy, and relatedness. See Medina-Gomez et al. for a detailed methods description of genetic methods.(Medina-Gomez et al., 2015)

In MAVAN, DNA was extracted from buccal epithelial cells and analyzed using the PsychChip and PsychArray (Illumina) genotyping platforms. Only SNPs shared across the two platforms were retained for further analysis. QC steps included sample call rate (retained above 95%), minor allele frequency (retained ≥1%), deviations from Hardy-Weinberg equilibrium (retained if $p<10^{-13}$), samples' heterozygosity rate, biological sex estimate, and cryptic relatedness (identity-by-descent) between samples. Complete method descriptions can be found in previous publications. (Garg et al., 2018; O'Donnell et al., 2014)

We included only children with European ancestry. In Generation R and ALSPAC non-European ancestry was defined by four or more standard deviations above or below the mean values of Hap Map European populations on any of the first four ancestry components in a multidimensional scaling analysis. In MAVAN ethnic outliers were defined as exceeding four standard deviations on any of the first four principal components relative to the mean MAVAN population, which is predominantly white (82%).

#### Child Psychopathology

The majority of measures were questionnaire-based, specifically the Child Behaviour Checklist (GenR, MAVAN),(Achenbach & Ruffle, 2000) Conners' Parent Rating Scale–Revised: Short Form (GenR, MAVAN),(Conners, Sitarenios, Parker, & Epstein, 1998) Social and Communication Disorders Checklist (ALSPAC),(Skuse, Mandy, & Scourfield, 2005) Social responsiveness scale (GenR),(Constantino, 2002) Strengths and Difficulties Questionnaire (ALSPAC, GenR, MAVAN)(Goodman, 1997) and the Pictorial Dominic Questionnaire (MAVAN)(Valla, Bergeron, & Smolla, 2000). Child self-rated measures included the Berkeley Puppet Interview (GenR)(Ringoot et al., 2013), and the Dominic (MAVAN), while diagnostic measures included the Development and Well-Being Assessment (ALSPAC)(Goodman, Ford, Richards, Gatward, & Meltzer, 2000) and the Preschool Age Psychiatric Assessment (MAVAN) (Egger et al., 2006).

#### Statistical Analysis

As the correlations between the PRSs was relatively modest (Figure S2) and thus multicollinearity was less of a concern than expected, we opted for a standard OLS regression approach using the optimal threshold as determined across all three cohorts, as opposed to an elastic net approach specified in the pre-study analysis plan.(Krapohl et al., 2018)

#### Attenuation of Depression Effects in Mutually Adjusted Models

We observed a substantial attenuation of regression estimates for the major depression and depressive symptoms PRS in the mutually adjusted model, compared to the separate PRS model. A potential explanation may be shared variance by the inclusion of two depression-related GWAS. We therefore performed an exploratory analysis with both depression PRSs included in the model, but no other PRS. The estimates were not substantially attenuated compared to the separate PRS model (Table S4). We therefore can conclude, that it is rather the combined inclusion of the other PRSs, which reduces the independent contribution of the depression PRSs

### Table S1

*Table 1: Variance explained in the general psychopathology factor by PRS in addition to covariates*

| | | ALSPAC (n=6575) | | GenR (n= 2418) | | MAVAN (n=254) | | Weighted Average (n=9247) |
|---|---|---|---|---|---|---|---|---|
| GWAS phenotype for PRS | PRS p cutoff | $n_{snps}$ | $\Delta R^2$ | $n_{snps}$ | $\Delta R^2$ | $n_{snps}$ | $\Delta R^2$ | avg. $\Delta R^2$ |
| Cognitive ability | 1 | 241478 | 1.44% | 254894 | 0.27% | 635450 | 0.02% | 0.99% |
| ADHD | 0.3 | 90150 | 1.21% | 92817 | 0.38% | 114615 | 2.31% | 0.98% |
| Major Depression | 0.05 | 37396 | 0.45% | 35609 | 0.15% | 36298 | 0.91% | 0.37% |
| Neuroticism | 0.01 | 16376 | 0.27% | 17140 | 0.42% | 33055 | 1.13% | 0.33% |
| Insomnia | 0.2 | 76321 | 0.40% | 73222 | 0.04% | 77273 | 0.04% | 0.26% |
| Schizophrenia | 0.05 | 40303 | 0.29% | 39055 | 0.21% | 47263 | 0.05% | 0.26% |
| Depressive symptoms | 0.3 | 85852 | 0.30% | 85067 | 0.00% | 99111 | 0.42% | 0.17% |
| Alcohol abuse | 0.2 | 96989 | 0.20% | 108774 | 0.07% | 126385 | 0.06% | 0.15% |
| Cross-disorder | 1 | 91597 | 0.12% | 108516 | 0.06% | 137243 | 1.07% | 0.11% |
| Bipolar | 0.0001 | 537 | 0.09% | 530 | 0.01% | 604 | 1.36% | 0.07% |
| Autism | 0.1 | 49864 | 0.03% | 46704 | 0.33% | 53252 | 0.34% | 0.09% |
| Total problems | 0.2 | 23912 | 0.06% | 24777 | 0.04% | 147656 | 0.10% | 0.06% |
| Generalized anxiety | 0.01 | 9290 | 0.04% | 8692 | 0.09% | 12641 | 0.01% | 0.05% |
| Social anxiety | 0.01 | 8559 | 0.07% | 7504 | 0.01% | 7372 | 0.61% | 0.06% |
| Panic | 0.00001 | 22 | 0.02% | 20 | 0.16% | 24 | 0.55% | 0.04% |
| Phobia | 0.2 | 101040 | 0.03% | 90089 | 0.03% | 93459 | 0.12% | 0.03% |
| Multi-PRS | 1 | 16 | 2.83% | 16 | 0.62% | 16 | -0.13% | 1.94% |
| Multi-PRS | 0.0005 | 7 | 2.81% | 7 | 0.64% | 7 | 0.46% | 2.02% |

**PRS p cutoff** P-value of PRS threshold with most significant association with outcome; for Multi-PRS model: indicator whether only PRS with significant association in separate PRS models (p<5E-04) were included
**$n_{snps}$** Number of SNPs included in PRS score; for Multi-PRS model: number of PRS included
**$\Delta R^2$** Variance explained in general psychopathology factor scores by PRS minus the variance explained by covariates. Multi-PRS $\Delta R^2$ is based on 10-fold cross-validation with 100 repetitions.
**avg. $\Delta R^2$** Sample size weighted average of $\Delta R^2$

### Table S2

*Table 2: Variance explained in the specific externalizing psychopathology factor by PRS in addition to covariates*

| | | ALSPAC (n=6575) | | GenR (n= 2418) | | MAVAN (n=254) | | Weighted Average (n=9247) |
|---|---|---|---|---|---|---|---|---|
| GWAS phenotype for PRS | PRS p cutoff | $n_{snps}$ | $\Delta R^2$ | $n_{snps}$ | $\Delta R^2$ | $n_{snps}$ | $\Delta R^2$ | avg. $\Delta R^2$ |
| ADHD | 0.5 | 122782 | 0.00% | 127335 | 1.74% | 157872 | 0.07% | 0.15% |
| Cognitive ability | 0.1 | 67618 | 0.03% | 69143 | 0.76% | 118885 | 0.51% | 0.13% |
| Autism | 0.4 | 135700 | 0.12% | 128647 | 0.16% | 147656 | 0.33% | 0.13% |
| Bipolar | 0.5 | 172528 | 0.11% | 184277 | 0.21% | 402261 | 0.07% | 0.13% |
| Total problems | 0.5 | 40801 | 0.01% | 43250 | 0.58% | 113431 | 0.33% | 0.08% |
| Cross-disorder | 0.5 | 63965 | 0.11% | 73569 | 0.03% | 94977 | 0.01% | 0.08% |
| Panic | 0.001 | 1244 | 0.02% | 1141 | 0.08% | 1364 | 0.21% | 0.03% |
| Major Depression | 0.05 | 37396 | 0.00% | 35609 | 0.49% | 36298 | 0.04% | 0.05% |
| Depressive symptoms | 0.3 | 85852 | 0.03% | 85067 | 0.26% | 99111 | 0.02% | 0.06% |
| Alcohol abuse | 0.00001 | 18 | 0.07% | 22 | 0.02% | 25 | 0.19% | 0.05% |
| Social anxiety | 0.01 | 8559 | 0.06% | 7504 | 0.03% | 7372 | 0.00% | 0.05% |
| Generalized anxiety | 0.05 | 35099 | 0.00% | 33265 | 0.10% | 52064 | 1.75% | 0.03% |
| Phobia | 0.00001 | 17 | 0.04% | 16 | 0.02% | 18 | 0.58% | 0.04% |
| Schizophrenia | 0.0001 | 1368 | 0.03% | 1383 | 0.01% | 1674 | 0.00% | 0.02% |
| Neuroticism | 0.001 | 4090 | 0.03% | 4173 | 0.00% | 6179 | 0.60% | 0.02% |
| Insomnia | 0.001 | 2392 | 0.04% | 2363 | 0.04% | 2530 | 0.00% | 0.04% |

**PRS p cutoff** P-value of PRS threshold with most significant association with outcome; for Multi-PRS model: indicator whether only PRS with significant association in separate PRS models (p<5E-04) were included
**$n_{snps}$** Number of SNPs included in PRS score; for Multi-PRS model: number of PRS included
**$\Delta R^2$** Variance explained in specific internalizing psychopathology factor scores by PRS minus the variance explained by covariates
**avg. $\Delta R^2$** Sample size weighted average of $\Delta R^2$

### Table S3

*Table 3: Variance explained in the specific internalizing psychopathology factor by PRS in addition to covariates*

| | | ALSPAC (n=6575) | | GenR (n= 2418) | | MAVAN (n=254) | | Weighted Average (n=9247) |
|---|---|---|---|---|---|---|---|---|
| GWAS phenotype for PRS | PRS p cutoff | $n_{snps}$ | $\Delta R^2$ | $n_{snps}$ | $\Delta R^2$ | $n_{snps}$ | $\Delta R^2$ | avg. $\Delta R^2$ |
| Neuroticism | 1 | 72208 | 0.42% | 81662 | 0.19% | 217279 | 1.99% | 0.38% |
| Generalized anxiety | 0.2 | 103464 | 0.17% | 99425 | 0.15% | 169260 | 0.00% | 0.16% |
| ADHD | 0.05 | 26334 | 0.08% | 26540 | 0.58% | 31922 | 0.96% | 0.18% |
| Cognitive ability | 0.0001 | 1636 | 0.05% | 1654 | 0.65% | 1686 | 0.11% | 0.15% |
| Schizophrenia | 0.4 | 149350 | 0.24% | 144192 | 0.00% | 172263 | 0.36% | 0.14% |
| Cross-disorder | 0.1 | 20947 | 0.19% | 22736 | 0.01% | 29232 | 0.68% | 0.14% |
| Major Depression | 0.2 | 95372 | 0.15% | 90624 | 0.07% | 95043 | 0.14% | 0.13% |
| Depressive symptoms | 0.3 | 85852 | 0.07% | 85067 | 0.32% | 99111 | 0.01% | 0.11% |
| Total problems | 1 | 52850 | 0.00% | 56546 | 0.37% | 147459 | 2.62% | 0.03% |
| Social anxiety | 1 | 267987 | 0.04% | 242884 | 0.18% | 254314 | 0.09% | 0.07% |
| Bipolar | 0.0001 | 537 | 0.13% | 530 | 0.01% | 604 | 0.13% | 0.09% |
| Phobia | 0.00001 | 17 | 0.07% | 16 | 0.00% | 18 | 1.28% | 0.05% |
| Autism | 0.01 | 8332 | 0.02% | 7789 | 0.13% | 8658 | 1.75% | 0.06% |
| Insomnia | 0.001 | 48673 | 0.02% | 46515 | 0.02% | 48932 | 1.75% | 0.02% |
| Panic | 0.5 | 192174 | 0.01% | 184423 | 0.07% | 275051 | 0.29% | 0.02% |
| Alcohol abuse | 0.0001 | 184 | 0.01% | 202 | 0.02% | 228 | 0.26% | 0.02% |

**PRS p cutoff** P-value of PRS threshold with most significant association with outcome; for Multi-PRS model: indicator whether only PRS with significant association in separate PRS models ($p<5E-04$) were included
$\mathbf{n_{snps}}$ Number of SNPs included in PRS score; for Multi-PRS model: number of PRS included
$\mathbf{\Delta R^2}$ Variance explained in specific internalizing psychopathology factor scores by PRS minus the variance explained by covariates
avg. $\Delta R^2$ Sample size weighted average of $\Delta R^2$

#### Table S4

*Table 4: General psychopathology factor regressed on depression PRS*

| GWAS phenotype for PRS | PRS p cutoff | ALSPAC (n=6575) β | ALSPAC (n=6575) SE | GenR (n= 2418) β | GenR (n= 2418) SE | MAVAN (n=254) β | MAVAN (n=254) SE |
|---|---|---|---|---|---|---|---|
| *Separate Models* | | | | | | | |
| Major Depression | 0.05 | 0.07 | 0.01 | 0.04 | 0.02 | 0.11 | 0.07 |
| Depressive symptoms | 0.3 | 0.05 | 0.01 | 0.00 | 0.02 | 0.07 | 0.07 |
| *Mutually adjusted for each other* | | | | | | | |
| Major Depression | 0.05 | 0.06 | 0.01 | 0.04 | 0.02 | 0.10 | 0.07 |
| Depressive symptoms | 0.3 | 0.04 | 0.01 | -0.01 | 0.02 | 0.05 | 0.07 |
| *Mutually adjusted for all PRS* | | | | | | | |
| Major Depression | 0.05 | 0.02 | 0.01 | 0.00 | 0.02 | 0.05 | 0.08 |
| Depressive symptoms | 0.3 | 0.02 | 0.01 | -0.03 | 0.02 | 0.03 | 0.08 |

**PRS p cutoff** P-value of PRS threshold with most significant association with outcome
**β** Standardized regression coefficient in standard deviations
**SE** Standard Error
**p** P-value of regression coefficient

### Figure S1

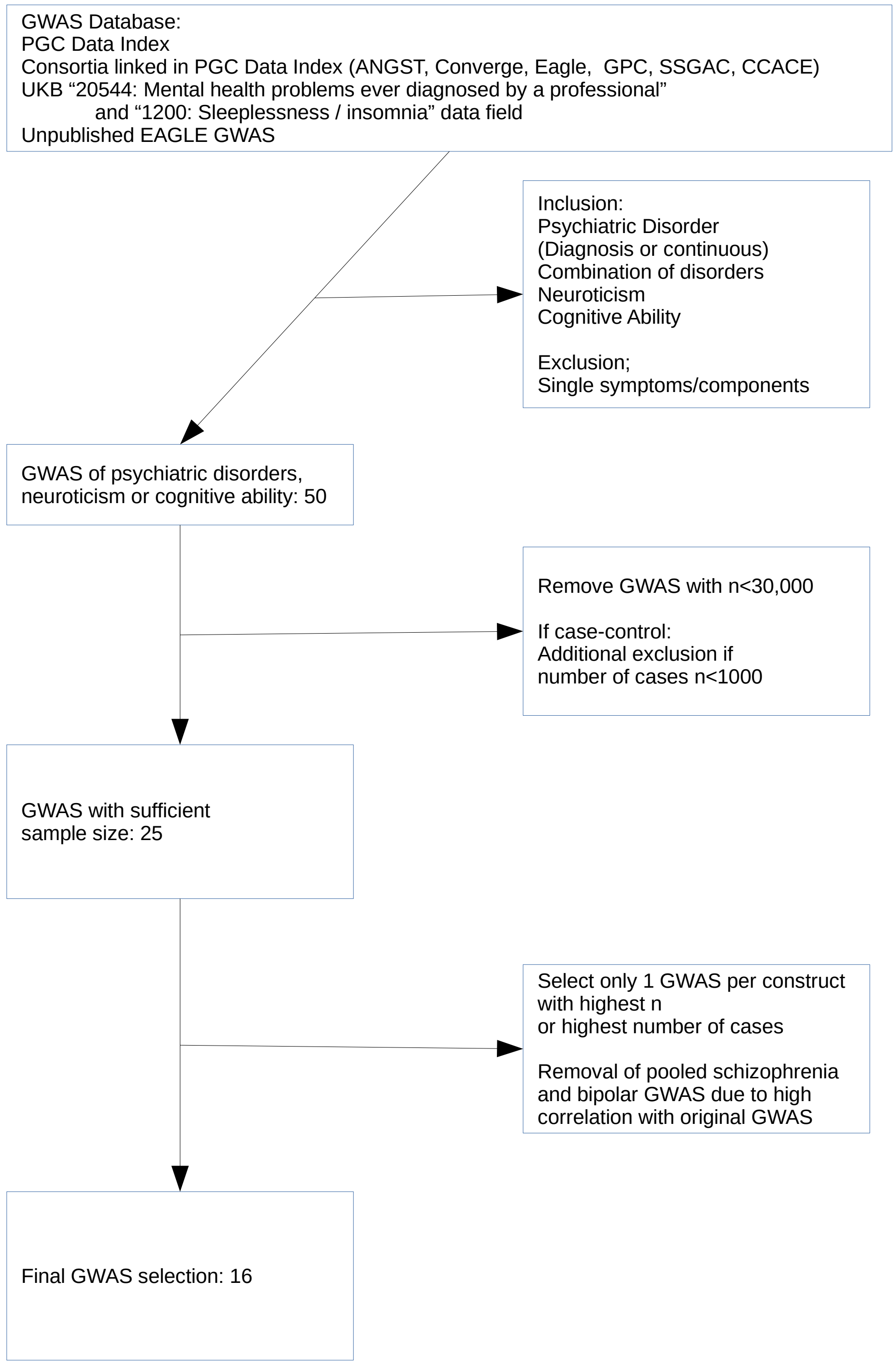
GWAS Database:
PGC Data Index
Consortia linked in PGC Data Index (ANGST, Converge, Eagle, GPC, SSGAC, CCACE)
UKB “20544: Mental health problems ever diagnosed by a professional”
and “1200: Sleeplessness / insomnia” data field
Unpublished EAGLE GWAS
Inclusion:
Psychiatric Disorder
(Diagnosis or continuous)
Combination of disorders
Neuroticism
Cognitive Ability
Exclusion;
Single symptoms/components
GWAS of psychiatric disorders,
neuroticism or cognitive ability: 50
Remove GWAS with n<30,000
If case-control:
Additional exclusion if
number of cases n<1000
GWAS with sufficient
sample size: 25
Select only 1 GWAS per construct
with highest n
or highest number of cases
Removal of pooled schizophrenia
and bipolar GWAS due to high
correlation with original GWAS
Final GWAS selection: 16

### Figure S2

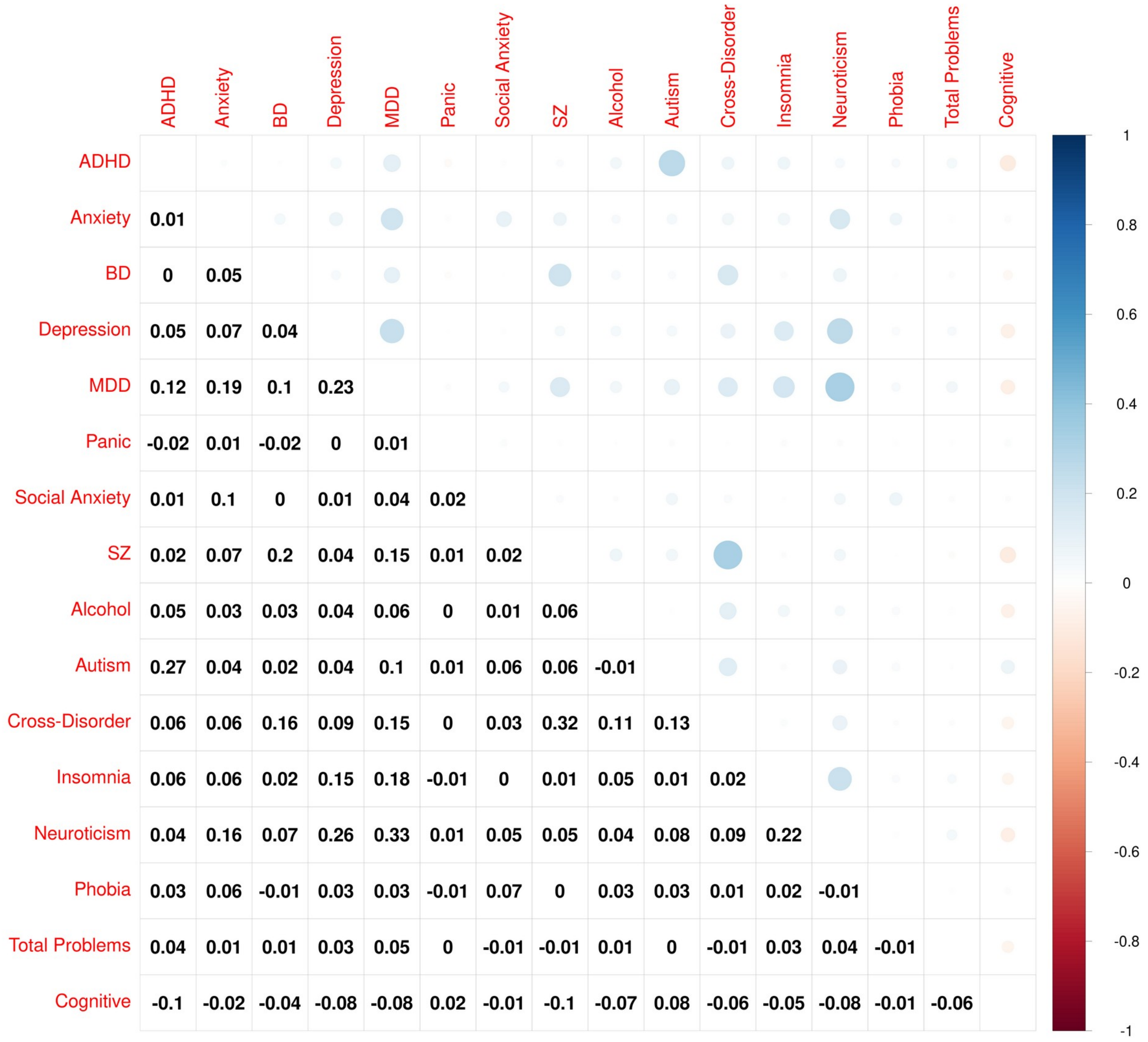


**Figure S2:** Correlation between the PRSs in the ALSPAC cohort at optimal threshold according to separate PRS model (n=6575).
